# COVID-19 individual participant data meta-analyses. Can there be too many? Results from a rapid systematic review

**DOI:** 10.1101/2022.09.01.22279485

**Authors:** Lauren Maxwell, Priya Shreedhar, Brooke Levis, Sayali Arvind Chavan, Shaila Akter, Mabel Carabali

## Abstract

**Background:** Individual participant data meta-analyses (IPD-MAs), which include harmonising and analysing participant-level data from related studies, provide several advantages over aggregate data meta-analyses, which pool study-level findings. IPD-MAs are especially important for building and evaluating diagnostic and prognostic models, making them an important tool for informing the research and public health responses to COVID-19.

**Methods:** We conducted a rapid systematic review of protocols and publications from planned, ongoing, or completed COVID-19-related IPD-MAs to identify areas of overlap and maximise data request and harmonisation efforts. We searched four databases using a combination of text and MeSH terms. Two independent reviewers determined eligibility at the title-abstract and full-text stage. Data were extracted by one reviewer into a pretested data extraction form and subsequently reviewed by a second reviewer. Data were analysed using a narrative synthesis approach. A formal risk of bias assessment was not conducted.

**Results:** We identified 31 COVID-19-related IPD-MAs, including five living IPD-MAs and ten IPD-MAs that limited their inference to published data (e.g., case reports). We found overlap in study designs, populations, exposures, and outcomes of interest. For example, 26 IPD-MAs included RCTs; 17 IPD-MAs were limited to hospitalised patients. Sixteen IPD-MAs focused on evaluating medical treatments, including six IPD-MAs for antivirals, four on antibodies, and two that evaluated convalescent plasma.

**Conclusions:** Collaboration across related IPD-MAs can leverage limited resources and expertise by expediting the creation of cross-study participant-level data datasets, which can, in turn, fast-track evidence synthesis for the improved diagnosis and treatment of COVID-19.

## BACKGROUND

The harmonisation and analysis of participant-level data and metadata for cross-study analyses, including individual participant data meta-analyses (IPD-MAs), can inform COVID-19 response through improved evaluation of diagnostic, preventative, and treatment measures. IPD-MAs have several analytic benefits over standard aggregate data meta-analyses when considering analyses of longitudinal data and the development and validation of clinical risk prediction tools.[1-3] IPD-MAs allow for joint consideration of study and subject-level heterogeneity to separate clinically relevant heterogeneity from heterogeneity related to study design or exposure and outcome ascertainment.[1-3] Separating clinically relevant from spurious heterogeneity is central to understanding whether observed differences in the risk of long COVID and COVID-19-related mortality are due to actual differences in exposure or immune response or due to study-level differences in selection, ascertainment, or residual confounding.

The implementation and management of IPD-MAs are resource-intensive.[1, 2, 4] Collecting the well-characterised metadata needed to appropriately describe included studies and cleaning and harmonising participant-level data from related studies require a significant investment of time and expertise from the primary studies and the IPD-MA management team.[2, 5] Additional barriers to sharing participant-level health-related data,[1] including fears of lost opportunities for publication and legal or ethical considerations, can prevent or slow down data sharing.[6-8] IPD-MAs are essential for informing research design, risk communication, and clinical practice for COVID-19. Given the significant resources needed to undertake an IPD-MA, identifying areas of overlap in exposures and outcomes of interest and inclusion criteria can foster cross-IPD-MA coordination to avoid duplication and maximise the utility of existing data.

## METHODS

We conducted a rapid systematic review to identify the protocols for or publications from planned, ongoing or completed COVID-19-related IPD-MAs. We described synergies across these efforts with a focus on study inclusion and exclusion criteria, including study population and study design, and exposure and outcomes of interest. We conducted a systematic search of four databases and protocol repositories, including Ovid Medline, the PROSPERO International Prospective Register of Systematic Reviews, the Open Science Foundation (OSF), and the Cochrane Database of Systematic Reviews, using a combination of MeSH (where applicable) and text terms (supplemental appendix table 1). We ran the searches on 2 June 2021, 29 October 2021, and 7 February 2022.

The protocol for this systematic review was developed per the Preferred Reporting Items for Systematic Reviews and Meta-Analyses (PRISMA)-Protocol statement guidelines.[9, 10] Before implementing the searches, we uploaded the systematic review protocol and search strategies to OSF (10.17605/OSF.IO/93GF2) after unsuccessfully trying to upload the protocol to the PROSPERO Registry of Systematic Reviews, which told our team that the systematic review of IPD-MAs was not a systematic review. This systematic review is reported in keeping with the 2020 PRISMA statement (supplemental appendix table 2).[11]

### Study selection and data extraction

Eligible protocols or published studies were IPD-MAs that planned to include or included participant-level COVID-19-related health data. IPD-MAs that only included social or psychological measures and systematic reviews that were limited to aggregate measures rather than participant-level data from included studies were excluded. Two independent reviewers determined eligibility at the title abstract and full-text screening stages. One reviewer extracted data into a pre-piloted data extraction Google sheet. Data were subsequently reviewed by a second reviewer. Differences of opinion and discrepancies in data extraction were resolved through consensus.

### Analysis

We conducted a narrative synthesis of the results and summarise findings in a series of Sankey diagrams created in RStudio 1.4.1103. We did not include a formal risk of bias assessment as part of this rapid systematic review, as most IPD-MAs only had a protocol available for review at the time of data extraction.

### Patient and public involvement

Patients and the public were not directly involved in this systematic review; we used publicly available data for the analysis.

## RESULTS

We reviewed 116 full texts and identified 31 COVID-19-focused health-related IPD-MAs (see supplemental appendix figure 1 for the PRISMA flow diagram). The majority of IPD-MAs were identified through PROSPERO (n=21), followed by Ovid Medline (n=8) and OSF (n=2).[12, 13] No IPD-MAs were identified from the Cochrane Database of Systematic Reviews. The 31 ongoing or completed COVID-19 IPD-MAs are described in Table 1. As shown in the Sankey diagrams in Figures 1A–D, there were several areas of overlap in included study populations, designs, interventions, and outcomes of interest between ongoing or completed and static or living COVID-19-related IPD-MAs. Figures 1C–D limit inference to the 21 IPD-MAs that requested data from authors, which requires more effort than IPD-MAs of data included in publications.

**Figure 1.**
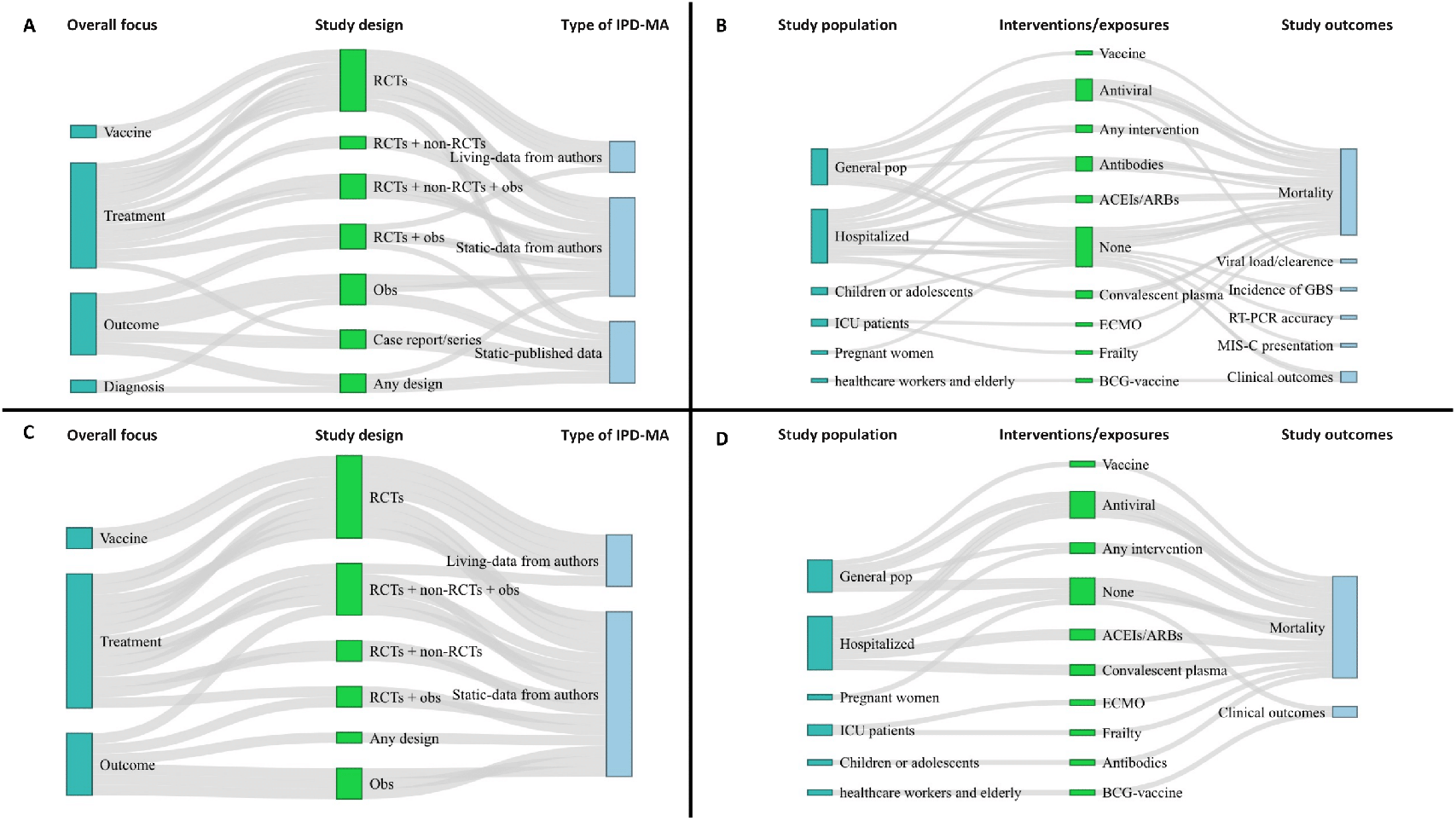
Sankey diagrams showing overlap between ongoing or completed and static or living COVID-19 IPD-MAs. **A**. Shows overlap between the focus, included study designs, and type of IPD-MA for all the ongoing or completed IPD-MAs. **B**. Shows overlap between the included study population, interventions/exposures, and outcomes of all the ongoing or completed IPD-MAs. **C**. Shows overlap between the focus, included study designs, and type of IPD-MA for only those IPD-MAs that requested data from authors. **D**. Shows overlap between the included study population, interventions/exposures, and outcomes of only those that requested data from authors. ACEIs=angiotensin-converting-enzyme inhibitors. ARBs=angiotensin II receptor blockers. BCG=Bacillus Calmette-Guérin. ECMO=extracorporeal membrane oxygenation. GBS=Guillain-Barré syndrome. MIS-C= multisystem inflammatory syndrome in children. Obs=observational. RCTs=randomised controlled trials. RT-PCR=reverse transcription polymerase chain reaction.

**Table 1.**
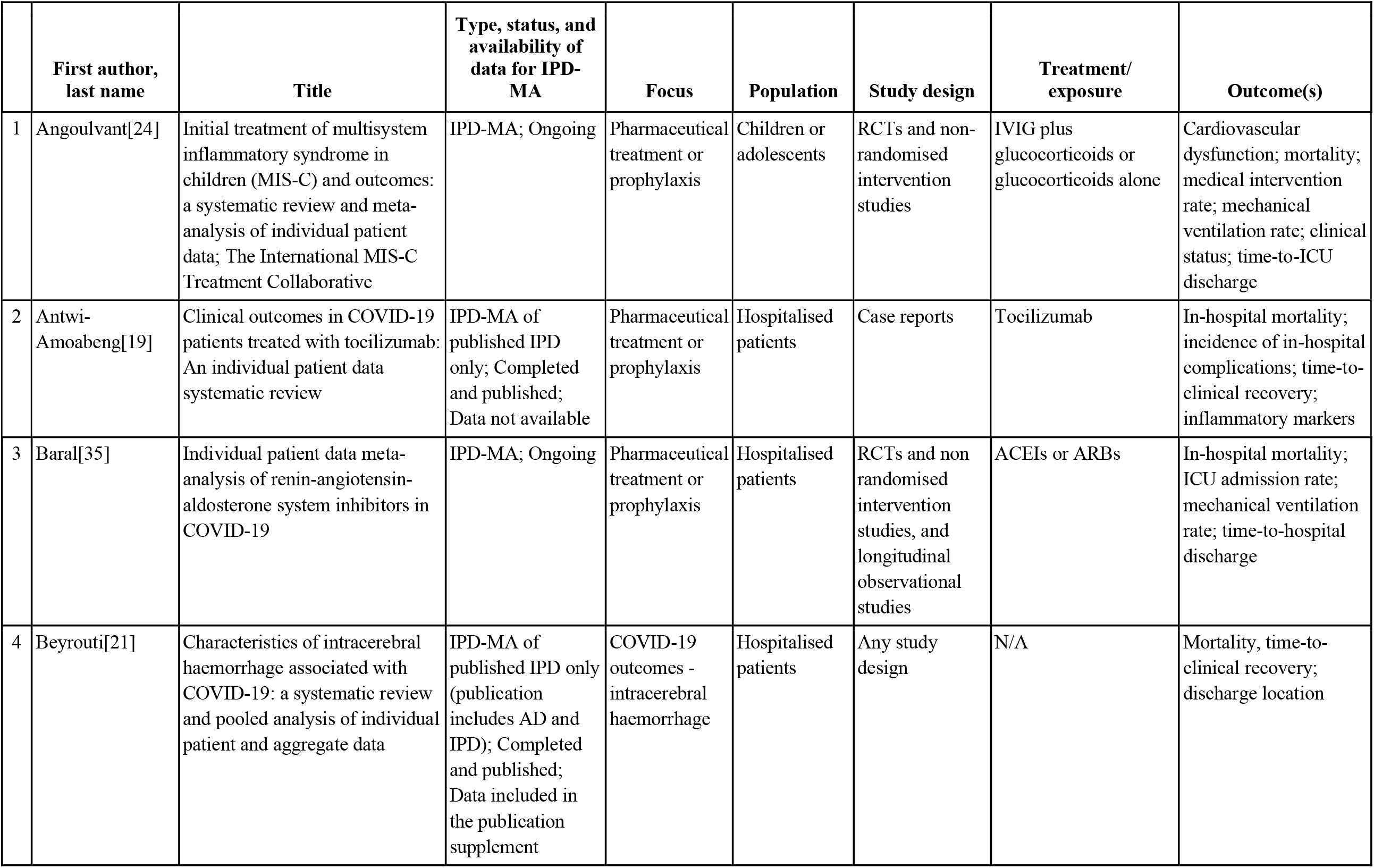
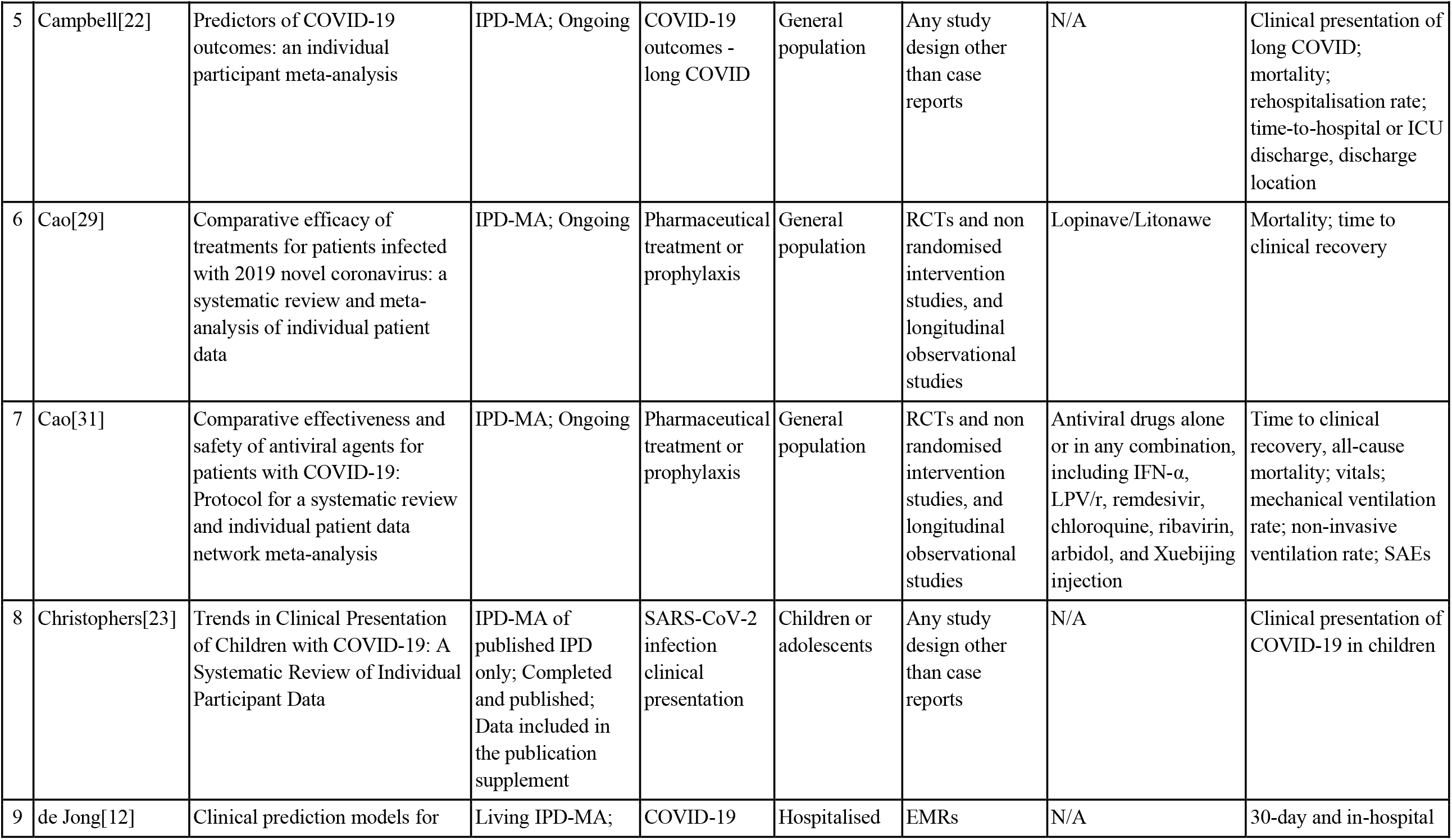
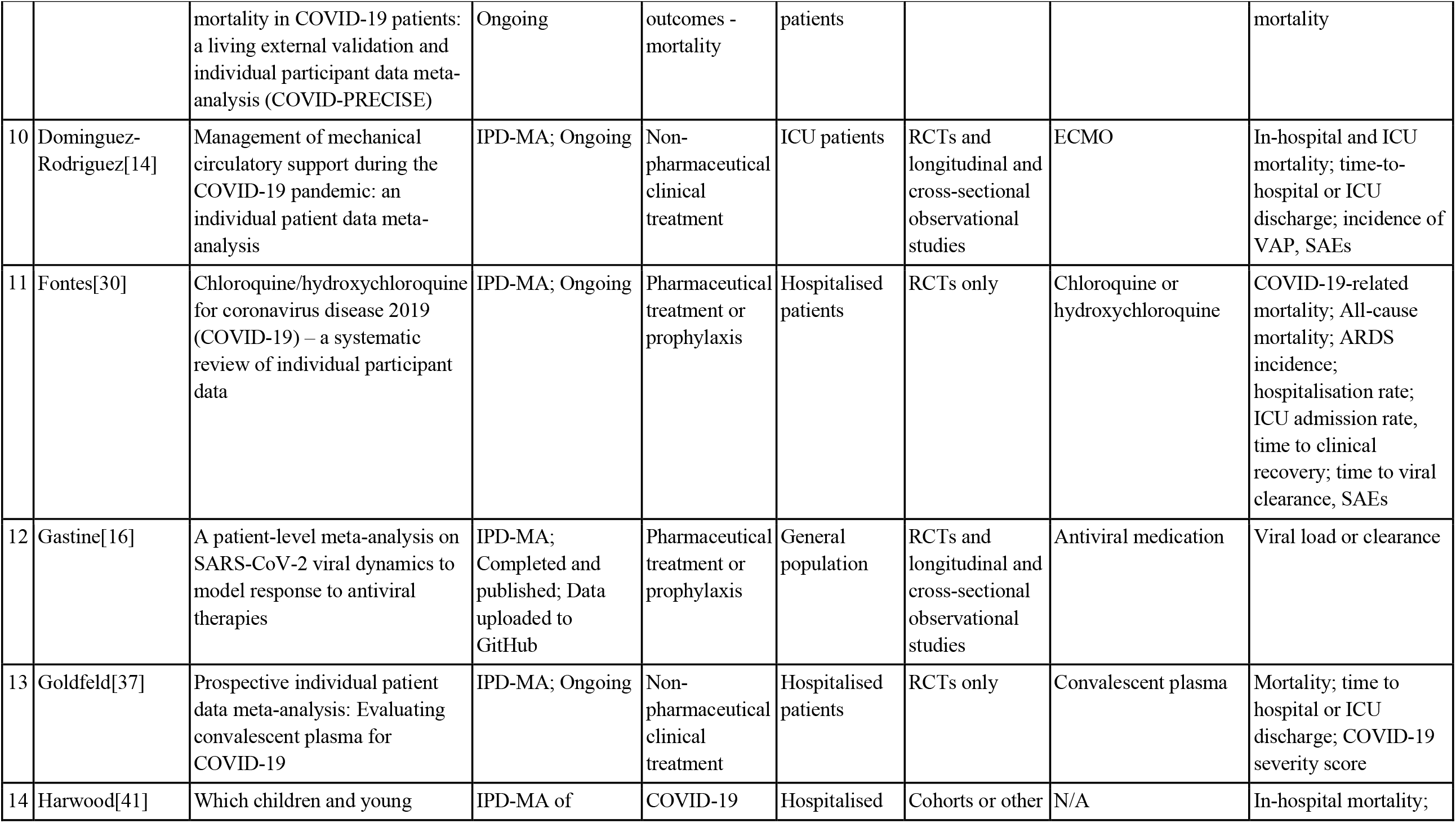
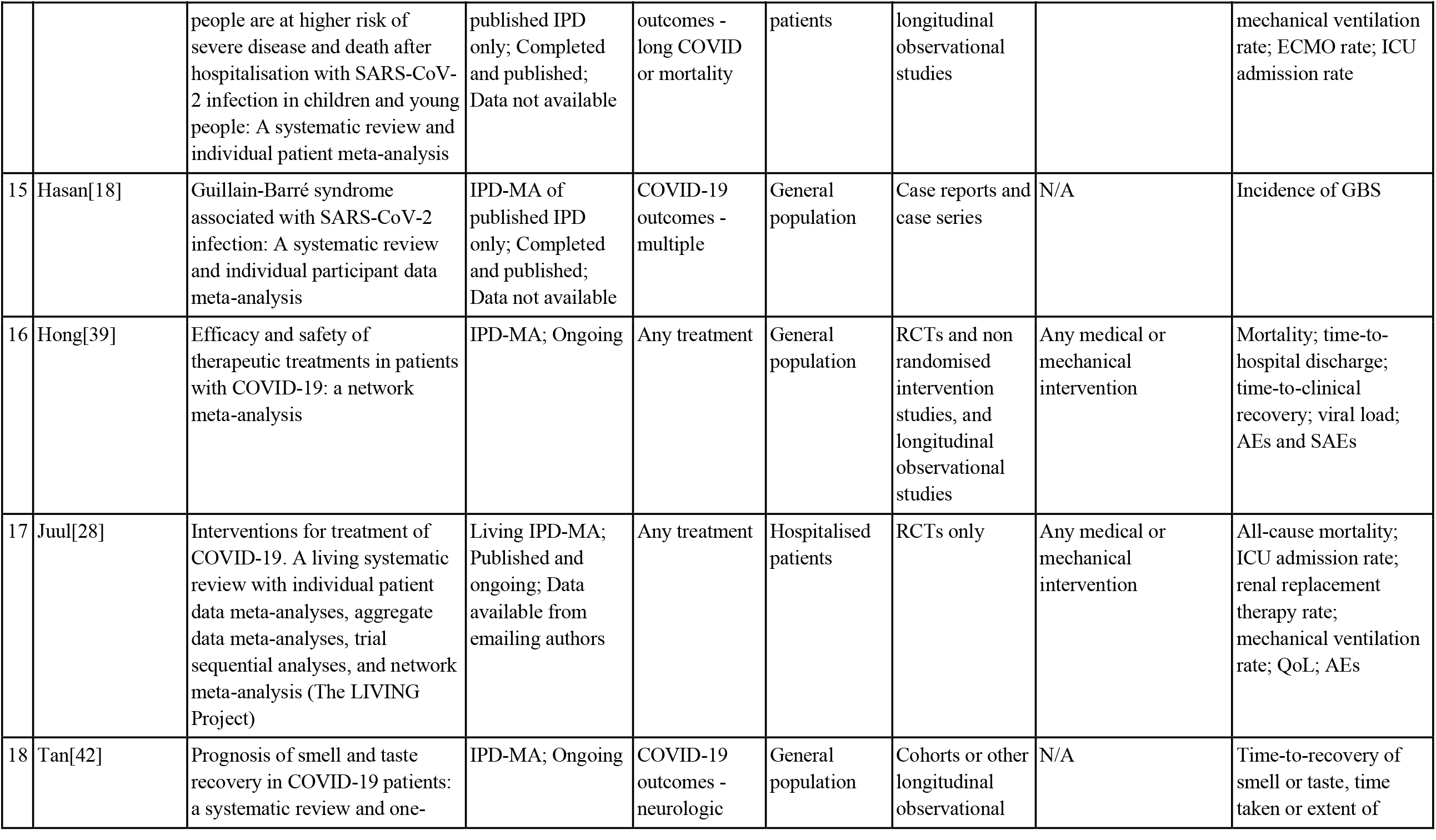
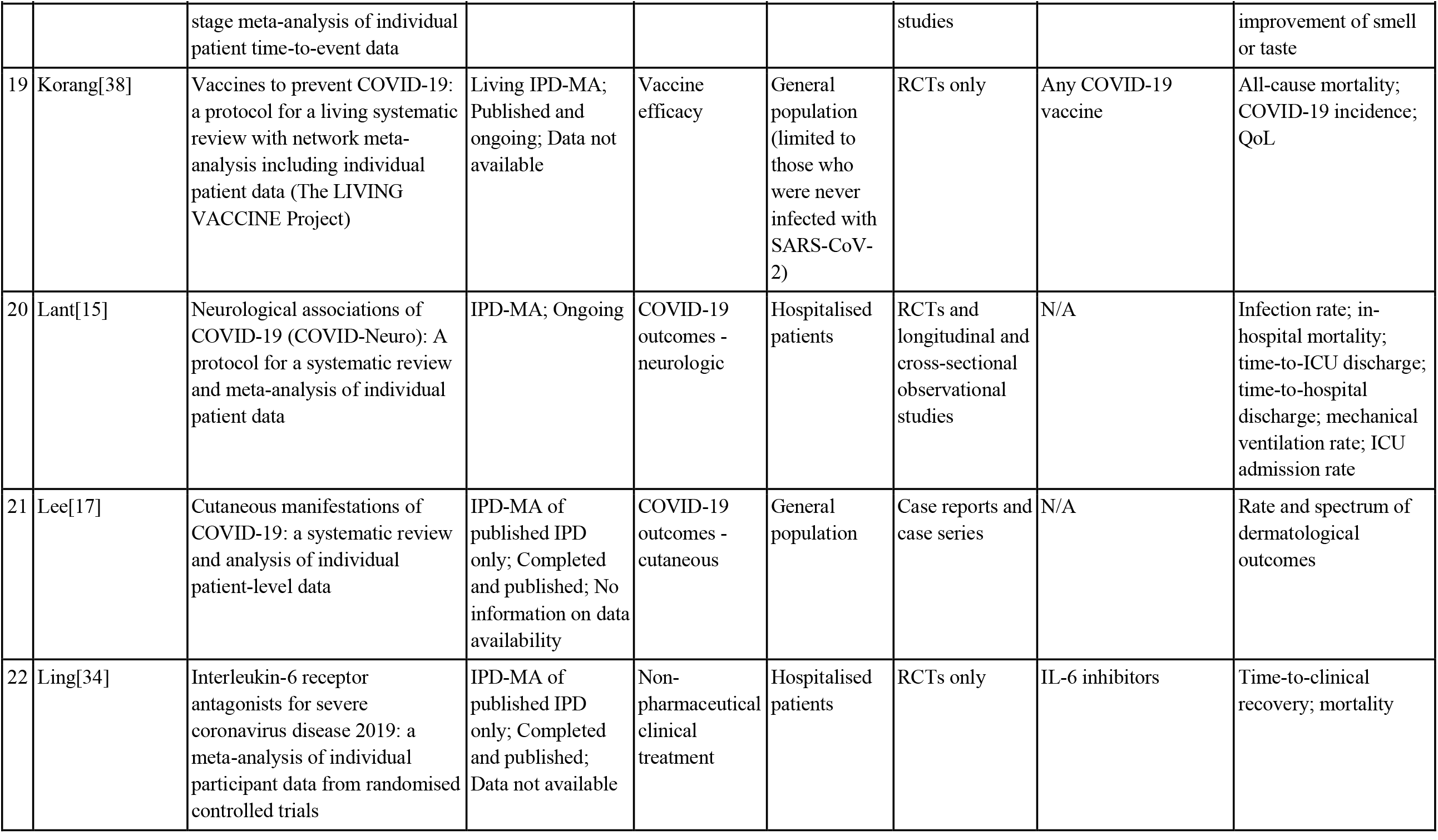
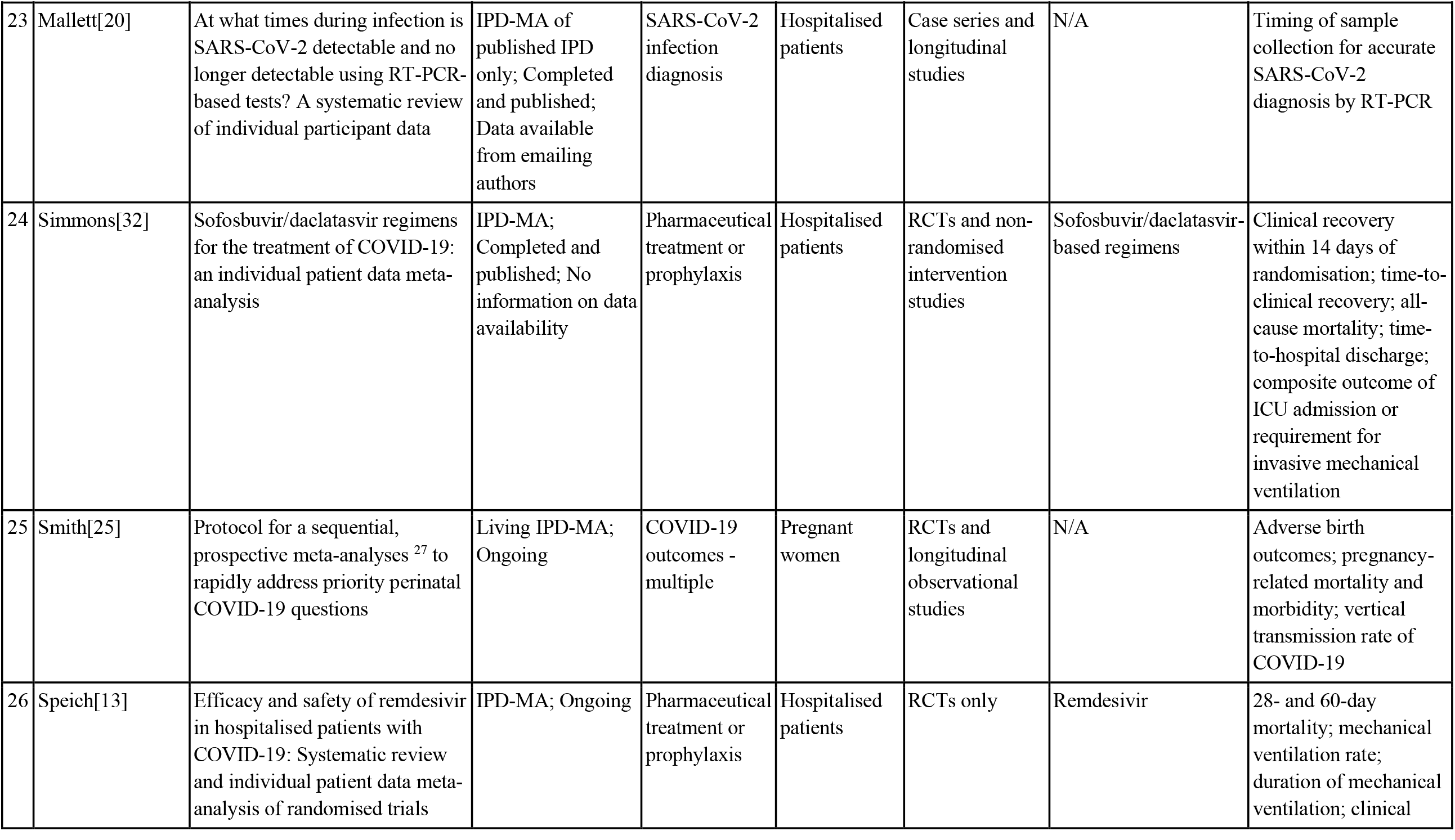
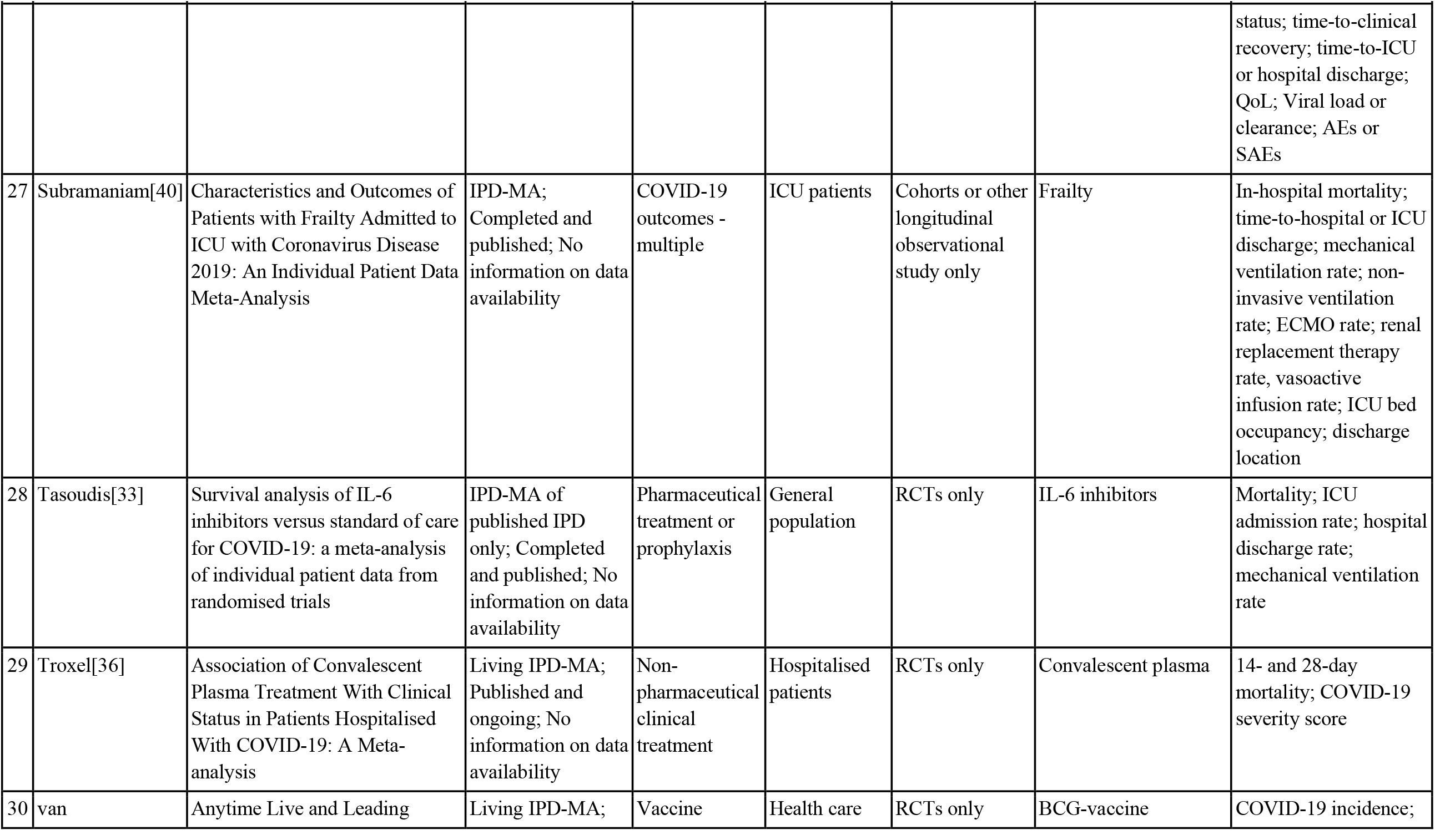
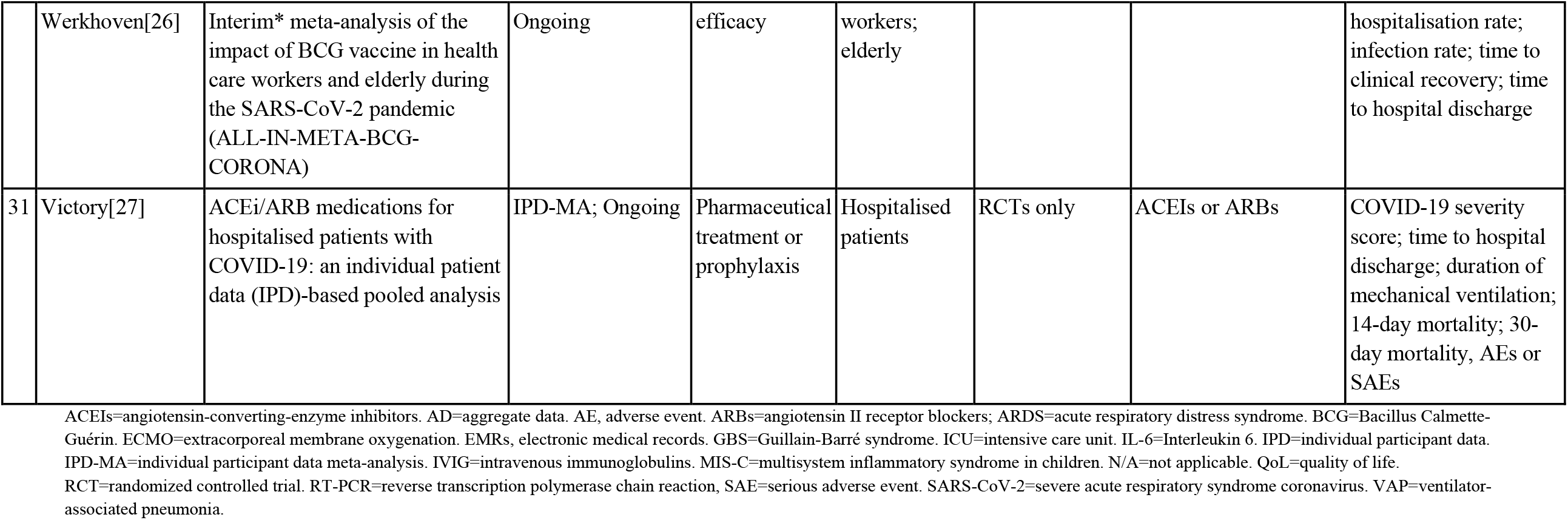
Overview of COVID-19-focused IPD-MAs

|  | **First author,**<br>**last name** | **Title** | **Type, status, and**<br>**availability of**<br>**data for IPD-**<br>**MA** | **Focus** | **Population** | **Study design** | **Treatment/**<br>**exposure** | **Outcome(s)** |
| --- | --- | --- | --- | --- | --- | --- | --- | --- |
| 1 | Angoulvant[24] | Initial treatment of multisystem inflammatory syndrome in children (MIS-C) and outcomes: a systematic review and meta-analysis of individual patient data; The International MIS-C Treatment Collaborative | IPD-MA; Ongoing | Pharmaceutical treatment or prophylaxis | Children or adolescents | RCTs and non-randomised intervention studies | IVIG plus glucocorticoids or glucocorticoids alone | Cardiovascular dysfunction; mortality; medical intervention rate; mechanical ventilation rate; clinical status; time-to-ICU discharge |
| 2 | Antwi-Amoabeng[19] | Clinical outcomes in COVID-19 patients treated with tocilizumab: An individual patient data systematic review | IPD-MA of published IPD only; Completed and published; Data not available | Pharmaceutical treatment or prophylaxis | Hospitalised patients | Case reports | Tocilizumab | In-hospital mortality; incidence of in-hospital complications; time-to-clinical recovery; inflammatory markers |
| 3 | Baral[35] | Individual patient data meta-analysis of renin-angiotensin-aldosterone system inhibitors in COVID-19 | IPD-MA; Ongoing | Pharmaceutical treatment or prophylaxis | Hospitalised patients | RCTs and non randomised intervention studies, and longitudinal observational studies | ACEIs or ARBs | In-hospital mortality; ICU admission rate; mechanical ventilation rate; time-to-hospital discharge |
| 4 | Beyrouti[21] | Characteristics of intracerebral haemorrhage associated with COVID-19: a systematic review and pooled analysis of individual patient and aggregate data | IPD-MA of published IPD only (publication includes AD and IPD); Completed and published; Data included in the publication supplement | COVID-19 outcomes - intracerebral haemorrhage | Hospitalised patients | Any study design | N/A | Mortality, time-to-clinical recovery; discharge location |
| 5 | Campbell[22] | Predictors of COVID-19 outcomes: an individual participant meta-analysis | IPD-MA; Ongoing | COVID-19 outcomes - long COVID | General population | Any study design other than case reports | N/A | Clinical presentation of long COVID; mortality; rehospitalisation rate; time-to-hospital or ICU discharge, discharge location |
| 6 | Cao[29] | Comparative efficacy of treatments for patients infected with 2019 novel coronavirus: a systematic review and meta-analysis of individual patient data | IPD-MA; Ongoing | Pharmaceutical treatment or prophylaxis | General population | RCTs and non randomised intervention studies, and longitudinal observational studies | Lopinave/Litonawe | Mortality; time to clinical recovery |
| 7 | Cao[31] | Comparative effectiveness and safety of antiviral agents for patients with COVID-19: Protocol for a systematic review and individual patient data network meta-analysis | IPD-MA; Ongoing | Pharmaceutical treatment or prophylaxis | General population | RCTs and non randomised intervention studies, and longitudinal observational studies | Antiviral drugs alone or in any combination, including IFN-α, LPV/r, remdesivir, chloroquine, ribavirin, arbidol, and Xuebijing injection | Time to clinical recovery, all-cause mortality; vitals; mechanical ventilation rate; non-invasive ventilation rate; SAEs |
| 8 | Christophers[23] | Trends in Clinical Presentation of Children with COVID-19: A Systematic Review of Individual Participant Data | IPD-MA of published IPD only; Completed and published; Data included in the publication supplement | SARS-CoV-2 infection clinical presentation | Children or adolescents | Any study design other than case reports | N/A | Clinical presentation of COVID-19 in children |
| 9 | de Jong[12] | Clinical prediction models for | Living IPD-MA; | COVID-19 | Hospitalised | EMRs | N/A | 30-day and in-hospital |

|  | First author,<br>last name | Title | Type, status, and<br>availability of<br>data for IPD-MA | Focus | Population | Study design | Treatment/<br>exposure | Outcome(s) |
| --- | --- | --- | --- | --- | --- | --- | --- | --- |
|  |  | mortality in COVID-19 patients: a living external validation and individual participant data meta-analysis (COVID-PRECISE) | Ongoing | outcomes - mortality | patients |  |  | mortality |
| 10 | Dominguez-Rodriguez[14] | Management of mechanical circulatory support during the COVID-19 pandemic: an individual patient data meta-analysis | IPD-MA; Ongoing | Non-pharmaceutical clinical treatment | ICU patients | RCTs and longitudinal and cross-sectional observational studies | ECMO | In-hospital and ICU mortality; time-to-hospital or ICU discharge; incidence of VAP, SAEs |
| 11 | Fontes[30] | Chloroquine/hydroxychloroquine for coronavirus disease 2019 (COVID-19) – a systematic review of individual participant data | IPD-MA; Ongoing | Pharmaceutical treatment or prophylaxis | Hospitalised patients | RCTs only | Chloroquine or hydroxychloroquine | COVID-19-related mortality; All-cause mortality; ARDS incidence; hospitalisation rate; ICU admission rate, time to clinical recovery; time to viral clearance, SAEs |
| 12 | Gastine[16] | A patient-level meta-analysis on SARS-CoV-2 viral dynamics to model response to antiviral therapies | IPD-MA; Completed and published; Data uploaded to GitHub | Pharmaceutical treatment or prophylaxis | General population | RCTs and longitudinal and cross-sectional observational studies | Antiviral medication | Viral load or clearance |
| 13 | Goldfeld[37] | Prospective individual patient data meta-analysis: Evaluating convalescent plasma for COVID-19 | IPD-MA; Ongoing | Non-pharmaceutical clinical treatment | Hospitalised patients | RCTs only | Convalescent plasma | Mortality; time to hospital or ICU discharge; COVID-19 severity score |
| 14 | Harwood[41] | Which children and young | IPD-MA of | COVID-19 | Hospitalised | Cohorts or other | N/A | In-hospital mortality; |
|  |  | people are at higher risk of severe disease and death after hospitalisation with SARS-CoV-2 infection in children and young people: A systematic review and individual patient meta-analysis | published IPD only; Completed and published; Data not available | outcomes - long COVID or mortality | patients | longitudinal observational studies |  | mechanical ventilation rate; ECMO rate; ICU admission rate |
| 15 | Hasan[18] | Guillain-Barré syndrome associated with SARS-CoV-2 infection: A systematic review and individual participant data meta-analysis | IPD-MA of published IPD only; Completed and published; Data not available | COVID-19 outcomes - multiple | General population | Case reports and case series | N/A | Incidence of GBS |
| 16 | Hong[39] | Efficacy and safety of therapeutic treatments in patients with COVID-19: a network meta-analysis | IPD-MA; Ongoing | Any treatment | General population | RCTs and non randomised intervention studies, and longitudinal observational studies | Any medical or mechanical intervention | Mortality; time-to-hospital discharge; time-to-clinical recovery; viral load; AEs and SAEs |
| 17 | Juul[28] | Interventions for treatment of COVID-19. A living systematic review with individual patient data meta-analyses, aggregate data meta-analyses, trial sequential analyses, and network meta-analysis (The LIVING Project) | Living IPD-MA; Published and ongoing; Data available from emailing authors | Any treatment | Hospitalised patients | RCTs only | Any medical or mechanical intervention | All-cause mortality; ICU admission rate; renal replacement therapy rate; mechanical ventilation rate; QoL; AEs |
| 18 | Tan[42] | Prognosis of smell and taste recovery in COVID-19 patients: a systematic review and one- | IPD-MA; Ongoing | COVID-19 outcomes - neurologic | General population | Cohorts or other longitudinal observational | N/A | Time-to-recovery of smell or taste, time taken or extent of |
|  |  | stage meta-analysis of individual patient time-to-event data |  |  |  | studies |  | improvement of smell or taste |
| 19 | Korang[38] | Vaccines to prevent COVID-19: a protocol for a living systematic review with network meta-analysis including individual patient data (The LIVING VACCINE Project) | Living IPD-MA; Published and ongoing; Data not available | Vaccine efficacy | General population (limited to those who were never infected with SARS-CoV-2) | RCTs only | Any COVID-19 vaccine | All-cause mortality; COVID-19 incidence; QoL |
| 20 | Lant[15] | Neurological associations of COVID-19 (COVID-Neuro): A protocol for a systematic review and meta-analysis of individual patient data | IPD-MA; Ongoing | COVID-19 outcomes - neurologic | Hospitalised patients | RCTs and longitudinal and cross-sectional observational studies | N/A | Infection rate; in-hospital mortality; time-to-ICU discharge; time-to-hospital discharge; mechanical ventilation rate; ICU admission rate |
| 21 | Lee[17] | Cutaneous manifestations of COVID-19: a systematic review and analysis of individual patient-level data | IPD-MA of published IPD only; Completed and published; No information on data availability | COVID-19 outcomes - cutaneous | General population | Case reports and case series | N/A | Rate and spectrum of dermatological outcomes |
| 22 | Ling[34] | Interleukin-6 receptor antagonists for severe coronavirus disease 2019: a meta-analysis of individual participant data from randomised controlled trials | IPD-MA of published IPD only; Completed and published; Data not available | Non-pharmaceutical clinical treatment | Hospitalised patients | RCTs only | IL-6 inhibitors | Time-to-clinical recovery; mortality |
| 23 | Mallett[20] | At what times during infection is SARS-CoV-2 detectable and no longer detectable using RT-PCR-based tests? A systematic review of individual participant data | IPD-MA of published IPD only; Completed and published; Data available from emailing authors | SARS-CoV-2 infection diagnosis | Hospitalised patients | Case series and longitudinal studies | N/A | Timing of sample collection for accurate SARS-CoV-2 diagnosis by RT-PCR |
| 24 | Simmons[32] | Sofosbuvir/daclatasvir regimens for the treatment of COVID-19: an individual patient data meta-analysis | IPD-MA; Completed and published; No information on data availability | Pharmaceutical treatment or prophylaxis | Hospitalised patients | RCTs and non-randomised intervention studies | Sofosbuvir/daclatasvir-based regimens | Clinical recovery within 14 days of randomisation; time-to-clinical recovery; all-cause mortality; time-to-hospital discharge; composite outcome of ICU admission or requirement for invasive mechanical ventilation |
| 25 | Smith[25] | Protocol for a sequential, prospective meta-analyses <sup>27</sup> to rapidly address priority perinatal COVID-19 questions | Living IPD-MA; Ongoing | COVID-19 outcomes - multiple | Pregnant women | RCTs and longitudinal observational studies | N/A | Adverse birth outcomes; pregnancy-related mortality and morbidity; vertical transmission rate of COVID-19 |
| 26 | Speich[13] | Efficacy and safety of remdesivir in hospitalised patients with COVID-19: Systematic review and individual patient data meta-analysis of randomised trials | IPD-MA; Ongoing | Pharmaceutical treatment or prophylaxis | Hospitalised patients | RCTs only | Remdesivir | 28- and 60-day mortality; mechanical ventilation rate; duration of mechanical ventilation; clinical |

|  | First author,<br>last name | Title | Type, status, and<br>availability of<br>data for IPD-MA | Focus | Population | Study design | Treatment/<br>exposure | Outcome(s) |
| --- | --- | --- | --- | --- | --- | --- | --- | --- |
|  |  |  |  |  |  |  |  | status; time-to-clinical recovery; time-to-ICU or hospital discharge; QoL; Viral load or clearance; AEs or SAEs |
| 27 | Subramaniam[40] | Characteristics and Outcomes of Patients with Frailty Admitted to ICU with Coronavirus Disease 2019: An Individual Patient Data Meta-Analysis | IPD-MA; Completed and published; No information on data availability | COVID-19 outcomes - multiple | ICU patients | Cohorts or other longitudinal observational study only | Frailty | In-hospital mortality; time-to-hospital or ICU discharge; mechanical ventilation rate; non-invasive ventilation rate; ECMO rate; renal replacement therapy rate, vasoactive infusion rate; ICU bed occupancy; discharge location |
| 28 | Tasoudis[33] | Survival analysis of IL-6 inhibitors versus standard of care for COVID-19: a meta-analysis of individual patient data from randomised trials | IPD-MA of published IPD only; Completed and published; No information on data availability | Pharmaceutical treatment or prophylaxis | General population | RCTs only | IL-6 inhibitors | Mortality; ICU admission rate; hospital discharge rate; mechanical ventilation rate |
| 29 | Troxel[36] | Association of Convalescent Plasma Treatment With Clinical Status in Patients Hospitalised With COVID-19: A Meta-analysis | Living IPD-MA; Published and ongoing; No information on data availability | Non-pharmaceutical clinical treatment | Hospitalised patients | RCTs only | Convalescent plasma | 14- and 28-day mortality; COVID-19 severity score |
| 30 | van | Anytime Live and Leading | Living IPD-MA; | Vaccine | Health care | RCTs only | BCG-vaccine | COVID-19 incidence; |
|  | Werkhoven[26] | Interim* meta-analysis of the impact of BCG vaccine in health care workers and elderly during the SARS-CoV-2 pandemic (ALL-IN-META-BCG-CORONA) | Ongoing | efficacy | workers;<br>elderly |  |  | hospitalisation rate; infection rate; time to clinical recovery; time to hospital discharge |
| 31 | Victory[27] | ACEi/ARB medications for hospitalised patients with COVID-19: an individual patient data (IPD)-based pooled analysis | IPD-MA; Ongoing | Pharmaceutical treatment or prophylaxis | Hospitalised patients | RCTs only | ACEIs or ARBs | COVID-19 severity score; time to hospital discharge; duration of mechanical ventilation; 14-day mortality; 30-day mortality, AEs or SAEs |

ACEIs=angiotensin-converting-enzyme inhibitors. AD=aggregate data. AE, adverse event. ARBs=angiotensin II receptor blockers; ARDS=acute respiratory distress syndrome. BCG=Bacillus Calmette-Guérin. ECMO=extracorporeal membrane oxygenation. EMRs, electronic medical records. GBS=Guillain-Barré syndrome. ICU=intensive care unit. IL-6=Interleukin 6. IPD=individual participant data. IPD-MA=individual participant data meta-analysis. IVIG=intravenous immunoglobulins. MIS-C=multisystem inflammatory syndrome in children. N/A=not applicable. QoL=quality of life. RCT=randomized controlled trial. RT-PCR=reverse transcription polymerase chain reaction, SAE=serious adverse event. SARS-CoV-2=severe acute respiratory syndrome coronavirus. VAP=ventilator-associated pneumonia.

### Study designs

Ten IPD-MAs included randomised controlled trials (RCTs), non-randomised intervention studies, or longitudinal observational studies; an additional 10 IPD-MAs were limited to RCTs only. Three IPD-MAs included RCTs and longitudinal or cross-sectional observational studies.[14-16] Two IPD-MAs had case reports and case series.[17, 18] One IPD-MA each was limited to case reports,[19] medical records,[12] and case series and longitudinal studies.[20] One IPD-MA included any study design;[21] two others included any study design other than case reports.[22, 23]

### Populations

More than half of the 31 IPD-MAs were conducted with data from hospitalised or intensive care unit (ICU) patients (n=17). Ten IPD-MAs included data from the general population, and two IPD-MAs were limited to children or adolescents.[23, 24] One IPD-MA was conducted with pregnant women[25] and one with older adults and health care workers.[26] Most IPD-MAs were not limited by geography (n=28). One IPD-MA was limited to studies in the US and Canada,[27] another to the US, Europe, and China,[28] and one to China.[29]

### Treatment or exposure

Sixteen IPD-MAs focused on the evaluation of medical treatments, including antivirals (n=6),[13, 16, 29-32] antibodies (n=4),[19, 24, 33, 34] angiotensin-converting-enzyme inhibitors (ACEIs) or angiotensin II receptor blockers (ARBs; n=2),[27, 35] convalescent plasma (n=2),[36, 37] COVID-19 vaccines (n=1),[38] and the Bacillus Calmette–Guérin (BCG)-vaccine (n=1).[26] One IPD-MA focused on extracorporeal membrane oxygenation (ECMO).[14] Two IPD-MAs evaluated any medical or mechanical intervention, including ECMO.[28, 39] One IPD-MA had frailty as the exposure.[40]

### Outcomes

IPD-MAs shared a number of common outcomes, including rate of mechanical (n=8) or non-invasive ventilation (n=2),[31, 40] ECMO rate (n=2),[40, 41] rate of serious adverse events (SAEs) or adverse events (AEs; n=7), viral clearance or viral load (n=4),[13, 16, 30, 39] COVID-19 infection rate (n=3),[15, 26, 38] rate of hospitalization or rehospitalization (n=3)[22, 26, 30] or admittance to the ICU (n=6), time-to-hospital or ICU discharge (n=12), hospital discharge location (n=3),[21, 22, 40] time-to-clinical recovery (n=10), COVID-19 severity score (n=3),[27, 36, 37] quality of life-related measures (n=3),[13, 28, 38] and mortality (n=24). Areas of overlap in mortality measures included IPD-MAs that assessed in-hospital mortality (n=7) and all-cause mortality (n=5). One IPD-MA assessed ICU mortality,[14] and another pregnancy-related mortality.[25] IPD-MAs that specified time-to-death, included: 14-day mortality (n=2),[27, 36] 28-day mortality (n=2),[13, 36] 30-day mortality (n=2),[12, 27] and 60-day mortality (n=1).[13] Individual IPD-MAs focused on the clinical presentation of long COVID-19[22] and COVID-19 in children;[23] time-to-recovery of smell or taste;[42] incidence of Guillain-Barre syndrome;[18] rate and spectrum of dermatological outcomes in COVID-19 patients;[17] viral load at of sample collection and its effect on the accuracy of RT-PCR;[20] and adverse birth outcomes and vertical transmission in pregnant women with COVID-19.[25]

### Types of IPD-MAs

Ten IPD-MAs were limited to published IPD, which means that the group conducting the IPD-MA did not contact authors to request data. Five were living IPD-MAs where datasets and related findings are regularly updated as evidence becomes available.[25, 26, 28, 36, 38] Living IPD-MAs included a real-time IPD-MA,[36] a network IPD-MA,[38] and an IPD-MA of IPD-MAs.[28] Living IPD-MAs focused on COVID-19 vaccines,[38] BCG vaccine,[26] any treatment,[28] convalescent plasma,[36] and issues of interest to perinatal populations.[25] Four of the five living IPD-MAs were limited to RCTs.[26, 28, 36, 38]

### Availability of data from IPD-MAs

Fifteen IPD-MAs were published when we submitted the manuscript for publication. Three published IPD-MAs made their data available through Github (n=1)[16] or the journal supplement (n=2).[21, 23] Two published IPD-MAs stated that interested researchers could request the dataset from the study team,[20, 28] five said that data would not be made available.[18, 19, 34, 38, 41] Five others did not include a statement related to data availability.[17, 32, 33, 36, 40] Three of the living IPD-MAs were published [28, 36, 38], although only one indicated that data could be requested from the study team.[28]

## DISCUSSION

IPD-MAs are an important tool for the rapid evidence generation needed to inform clinical practice, making them a vital part of the research response to emerging pathogens.[43] We conducted a rapid systematic review to identify ongoing or completed COVID-19-related IPD-MAs. There were many areas of overlap in the 31 COVID-19-related IPD-MAs, including in study design and population, exposure, and outcomes of interest. In particular, the 14 IPD-MAs that evaluated the same medical exposures (antivirals, antibodies, ACEIs and ARBs, and convalescent plasma) represent a missed opportunity to exploit synergies. Most IPD-MA protocols were registered on PROSPERO, which could flag these areas of overlap when researchers submit their protocol. IPD-MAs require a significant investment of time and expertise, both from the team conducting the IPD-MA and the groups contributing data to the IPD-MA. Rapidly identifying and exploiting shared inclusion criteria can help facilitate evidence generation and avoid unnecessary duplication of effort.

We identified at least 10 IPD-MAs that limited their analysis to data included in published reports. While IPD-MAs that are limited to published IPD have been conducted previously, the volume of the research response to COVID-19 coupled with the push for reproducibility and transparency have likely facilitated the rise in IPD-MAs of data that were included in the study publications. Almost half of the IPD-MAs of published data included case study or case series data (n=4/10; 40%).[17-20] Given that the utility of the IPD-MA is limited by the quality of the studies that contribute data,[2] findings from these rapidly produced IPD-MAs should be considered preliminary and updated when more detailed and less selective participant-level datasets become available. This finding is in keeping with a methodological review of published data that compared the methodological and reporting quality of COVID-19 and non-pandemic research and found a reduction in quality in the former.[44]

While we reviewed the protocols for all IPD-MAs, we could only identify the restriction to published IPD for those IPD-MAs that had published their analyses, which suggests a need to clarify inclusion criteria in IPD-MA protocols to specify the intent to limit inference to published IPD. Some of the unpublished studies identified in our review may be misclassified as having the classical approach to conducting an IPD-MA, which includes the challenges associated with requesting the data from the data producers.

Living IPD-MAs are regularly updated as more evidence becomes available, representing substantial investments. There was overlap in study design, exposure, and outcome measurements in several of the five living IPD-MAs and between the living IPD-MAs and static IPD-MAs, which represents an opportunity to share limited resources and expedite findings.

Only a few IPD-MAs of data received from authors had been published when this manuscript was submitted for publication (n=5/21; 24%), so we could not quantify the overlap in datasets across IPD-MAs that collected datasets from research teams which would be an important measure of cross-IPD-MA redundancy in efforts. Only three of the ten published IPD-MAs had made data available through a repository or the publication of supplementary materials,[16, 21, 23] which suggests a continued need to encourage data sharing.

Working collaboratively to harmonise and share data across related IPD-MAs would maximise limited resources and shorten the timeline to deliver results that best inform clinical and public health practice. Testing the same hypotheses, especially with the same study designs or populations, represents a missed opportunity to evaluate novel hypotheses. Our findings support similar calls from a living review of COVID-19-related clinical trials and a scoping review of COVID-19-related data sharing platforms, which urged coordination across initiatives to reduce redundancies.[45] We propose the creation of a task force to identify concrete steps to enable cross-initiative collaboration and ensure that the harmonised participant-level data and study-related metadata correspond to the findable, accessible, interoperable, reusable (FAIR) principles for data resources.[46] These steps could include a cross-platform algorithm that uses natural language processing to alert researchers to similar initiatives at the time of protocol deposition.

We have listed additional resources facilitate the design and conduct of an IPD-MA in the context of an emerging pathogen in Table 2. The global scope and rapidly evolving nature of the pandemic underscore the need for more meta-collaborations that work to bring together data sharing efforts and cross-national analyses. The coordination of ongoing or planned IPD-MAs is a good starting place.

**Table 2.**
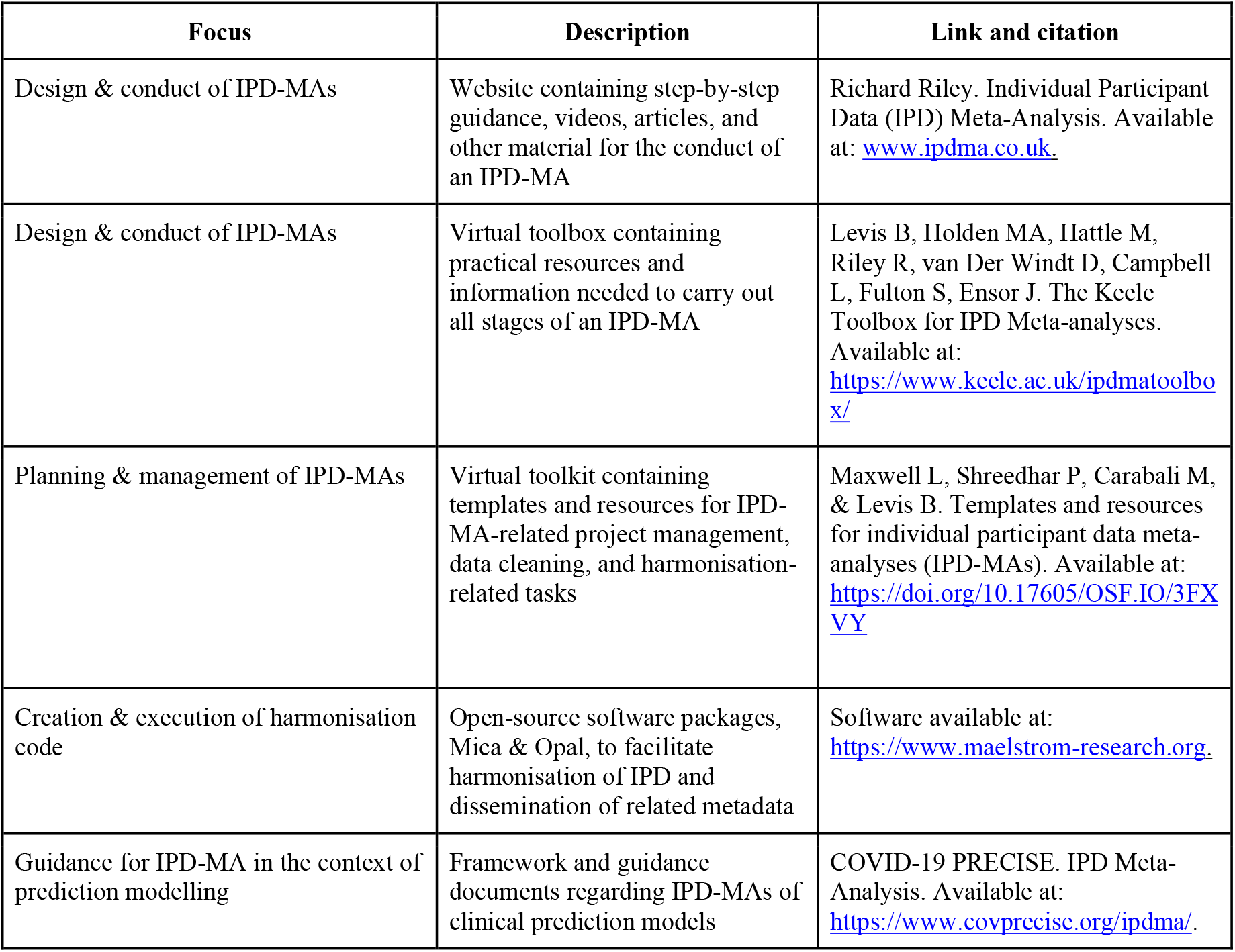
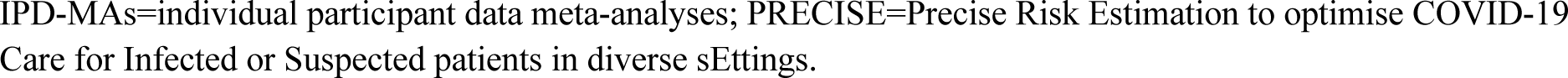
Resources for the conduct of an IPD-MA of an emerging pathogen IPD-MAs=individual participant data meta-analyses; PRECISE=Precise Risk Estimation to optimise COVID-19 Care for Infected or Suspected patients in diverse sEttings.

## CONCLUSION

IPD-MAs are important for informed research and public health response to COVID-19. To identify areas of overlap, we conducted a rapid systematic review of completed or ongoing COVID-19 IPD-MAs. We identified 31 COVID-19-related IPD-MAs, including five living IPD-MAs, and found several areas of overlap in study designs, populations, exposures, and outcomes of interest. This review shows several potential areas of collaboration across related IPD-MAs which can leverage limited resources and expertise by expediting the creation of cross-study participant-level datasets. This, in turn, can fast-track evidence synthesis for the improved diagnosis and treatment of COVID-19.

## Data Availability

A spreadsheet with comprehensive information on all planned or concluded IPD-MAs described in this review is available on Zenodo (10.5281/zenodo.6623480) under the Creative Commons Attribution 4.0 International (CC BY 4.0) license.

https://doi.org/10.5281/zenodo.6623480

## CONTRIBUTORS

LM, BL, and MC conceived of and designed the study. LM wrote the research protocol and developed the search strategy. LM, SA, and SC conducted the title abstract and full-text screening. LM, MC, PS, SA, and SC extracted and interpreted the data. LM wrote the first draft of the manuscript. All authors provided critical reviews of the manuscript. All authors had full access to all study data, take responsibility for data integrity and reliability of the analysis, and had final responsibility for the decision to submit for publication.

## COMPETING INTERESTS

None declared.

## FUNDING

ReCoDID project, funded by the EU Horizon 2020 research and innovation programme (grant agreement 825746) and the CIHR Institute of Genetics (grant agreement 01886-000) grant to LM.

## ROLE OF THE FUNDING SOURCE

The study’s funders had no role in the study design, data collection, data analysis, data interpretation, or report writing. The corresponding author had full access to all the data in the study and had final responsibility for the decision to submit this manuscript for publication.

## Notes

### Competing Interest Statement

The authors have declared no competing interest.

### Funding Statement

This study is funded by the ReCoDID project, funded by the EU Horizon 2020 research and innovation programme (grant agreement 825746), and the CIHR Institute of Genetics (grant agreement 01886-000) grant to Lauren Maxwell. The study's funders had no role in the study design, data collection, data analysis, data interpretation, or report writing.

